# CD19+ B cell numbers predict the increase of anti-SARS CoV2 antibodies in fingolimod-treated and COVID-19-vaccinated patients with multiple sclerosis

**DOI:** 10.1101/2022.07.02.22277178

**Authors:** I. Schiavetti, L. Barcellini, C. Lapucci, F. Tazza, M. Cellerino, E. Capello, D. Franciotta, M. Inglese, M. P. Sormani, A. Uccelli, A. Laroni

## Abstract

Treatment with fingolimod for multiple sclerosis (MS) reduces the efficacy of COVID-19 vaccination. We evaluated by a multivariate linear regression model whether main lymphocyte subsets and demographic feature correlated to the subsequent increase in anti-SARS-CoV2 antibodies following the third dose of COVID-19 vaccination in fingolimod-treated MS patients. We found that number and proportion of peripheral blood CD19+ B lymphocytes before the third dose of vaccination in MS patients treated with fingolimod, predict the subsequent increase of anti-SARS-CoV2 antibodies (respectively p = 0.013; p = 0.015). This work suggests that evaluating the numbers of CD19+ B cells may be important to identify patients at risk of not producing SARS-CoV-2 antibodies, with possible reduced protection from COVID-19.

## Introduction

COVID-19 vaccination is less effective in patients with multiple sclerosis (MS) treated with fingolimod, as evaluated through antibody response, T-cell specific responses and risk of developing breakthrough Covid-19. Even after the third dose of COVID-19 vaccine, only a fraction of patients treated with fingolimod develop an antibody response, whose mean levels are lower compared to untreated subjects^1^. The reason of the lower response to COVID-19 vaccine is only partially explained by the mechanism of action of fingolimod, a functional antagonist of the sphingosine-1-phosphare receptors, which causes a decrease of circulating T- and B-lymphocytes due to impaired recirculation. Total lymphopenia is the only factor which has been associated to impaired humoral response to COVID-19 vaccination in patients treated with fingolimod ^1^. The objective of the present study was to ascertain whether the numbers and proportion of circulating main lymphocyte subsets predict the antibody response to COVID-19 vaccination in MS patients treated with fingolimod.

## Methods

### Patients

This is a prospective single-center observational study including a subgroup of adult patients with MS (pwMS) in treatment with fingolimod just enrolled for CovaXiMS study. All of them underwent COVID-19 vaccination and were willing to provide blood samples for assessment of antibody levels and for the evaluation of immunological status before and after the three vaccination doses. Additional details about inclusion criteria are available elsewhere ^2^. All participants signed written informed consent before starting any study procedures. The study was conducted in accordance with all national regulations and with principles of the Declaration of Helsinki and the protocol was approved by the regional ethic committee (CER Liguria: 5/2021 - DB id 11169-21/01/2021) and at national level by AIFA/Spallanzani (Parere n 351, 2020/21).

### SARS CoV2 antibodies

Levels of anti-SARS-CoV-2 antibodies were measured within one month before the first dose of vaccine, one month after the second dose and within one month before and one month after the third dose, by a centralized laboratory with a double-antigen sandwich-based electrochemiluminescence immunoassay (ECLIA), using a commercial kit (Elecsys, Anti-SARS-CoV-2 S, Roche Diagnostics).

### Lymphocyte subsets

Total numbers of leukocytes, and total number and percentage of lymphocytes, CD3+ T lymphocytes, CD3+CD4+ lymphocytes, CD3+CD8+ T lymphocytes, CD19+ B lymphocytes, CD19+CD27+ memory B lymphocytes were measured through clinical immune profiling at the IRCCS Ospedale Policlinico San Martino before the third dose of COVID-19 vaccination.

### Statistical Analysis

The association of the antibody levels (reported natural logarithm of the difference between the post and pre third dose levels plus one) with age, sex, and each subset of lymphocytes (total number and percentage) was assessed by a multivariate linear regression model.

## Results

From January 2021 until April 2021, 20 adult pwMS on fingolimod (15 females/5 males) with a mean age of 49.3 ± 10.41 years were enrolled for the study. Fourteen (70.0%) had relapsing-remitting MS, median EDSS was 3.5 (IQR: 1.8 – 4.8), mean disease duration was 15.2 ± 11.38 years, and 75.0% had already been treated with different DMTs before fingolimod.

The only lymphocyte class significantly associated with a higher increase in the antibody level at multivariable analysis was the level of CD19+ lymphocytes (absolute number and percentage) (Figure 1), with an average 19% increase in antibody levels for each additional percentage increase in CD19+ lymphocytes (Table 1A) and an average 1% increase in antibody levels for each additional number/mm^3^ of CD19+ lymphocyte (Table 1B). For both analysis increasing age was significantly associated to a lower level of antibodies.

**Figure 1:**
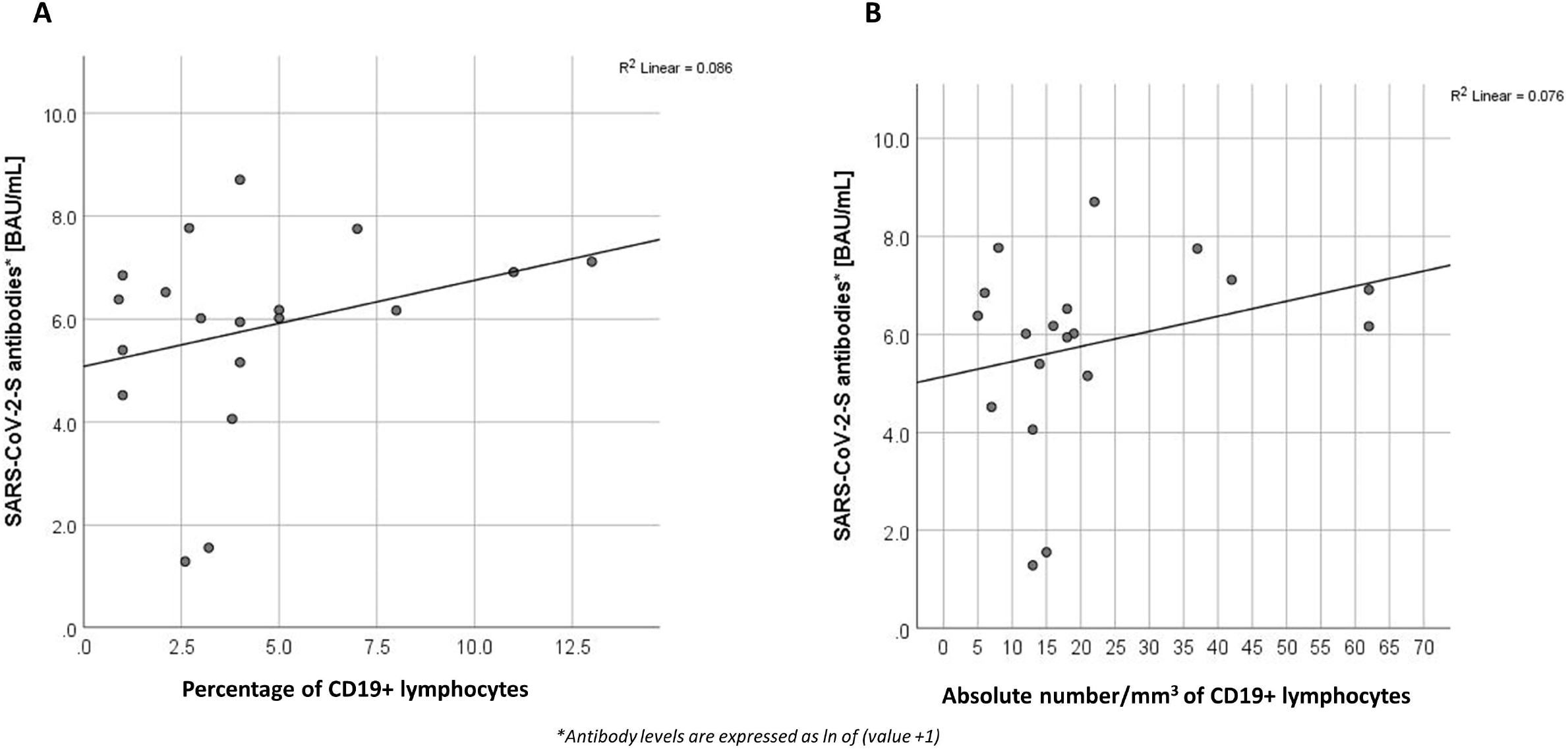
Correlation between level of CD19+ lymphocytes (1A: percentage; 1B: absolute number) and antibody level after the third vaccination dose.

**Table 1 A.** Factors associated to antibody levels, after the third vaccination dose.

| Parameter | Beta coefficients (SE) | Sig. |
| --- | --- | --- |
| Sex (females vs males) | 0.018 (0.841) | 0.98 |
| Age | - 0.092 (0.033) | 0.006 |
| Percentage of CD19+ lymphocytes | 0.188 (0.077) | 0.015 |

*Dependent Variable: $\ln(\Delta\text{Post-Pre third dose} + 1)$*
*Model: (Intercept), Sex, Age, Percentage of CD19+ lymphocytes*

**Table 1 B.** Factors associated to antibody levels, after the third vaccination dose.

| Parameter | Beta coefficients (SE) | Sig. |
| --- | --- | --- |
| Sex (females vs males) | 0.347 (0.797) | 0.66 |
| Age | - 0.100 (0.033) | 0.003 |
| Absolute numbers/mm<sup>3</sup> of CD19+ lymphocytes | 0.013 (0.005) | 0.013 |

*Dependent Variable: $\ln(\Delta\text{Post-Pre third dose} + 1)$*
*Model: (Intercept), Sex, Age, Absolute numbers/mm<sup>3</sup> of CD19+ lymphocytes*

## Discussion

The results of this study demonstrate that the number and proportion of peripheral blood CD19+ B lymphocytes before the third dose of COVID-19 vaccination, in patients treated with fingolimod, predict the subsequent increase of anti-SARS CoV2 antibodies.

Low or undetectable levels of specific antibodies after COVID-19 vaccination are a concern, since, as we have previously reported, every 10x increase in the antibody concentration reduces by 49% the risk of breakthrough COVID-19 ^3^. Achiron and colleagues observed a lower rate of seroconversion following the third dose of COVID-19 vaccination in patients who continued fingolimod compared to those who suspended and received the third dose after the total number of lymphocytes was 1000/mm3 ^1^.

In this cohort, total number of lymphocytes, or T lymphocytes did not correlate with changes in the antibody levels. This may be at least in part surprising, since previous work in the animal model has suggested that fingolimod affects production of antibodies in a T-cell dependent fashion ^4^. On the contrary, we found that the number and proportion of CD19+ B cells were associated to the increase of specific antibodies following the third dose of vaccine. Besides the number of CD19+ B cells, higher age was associated to lower development of antibodies, confirming that senescence of the immune system is linked to impaired response to vaccination. Similarly, CD19+ levels and age, together with IgM levels, were recently reported to predict the humoral response to anti-SARS CoV2 vaccination in patients with hematological malignancies ^5^.

This study has some limitations: first of all, the low number of enrolled subjects, secondly the fact that immune cell subsets before the first dose of vaccine were not available.

If confirmed by other studies, the results of this study suggest that assessing the numbers of CD19+ B cells in fingolimod-treated patients may be useful for identifying patients at higher risk of responding poorly to COVID-19 vaccination.

## Data Availability

All data produced in the present study are available upon reasonable request to the authors

## Funding

This work was supported by grants from Italian Ministry of Health (Ricerca Corrente). -This work was developed within the framework of the DINOGMI Department of Excellence of MIUR 2018-2022 (legge 232 del 2016). The CovaXiMS study was supported by FISM - Fondazione Italiana Sclerosi Multipla - cod. 2021/Special-Multi/001 and financed or co-financed with the ‘5 per mille’ public funding.

## Conflict of Interest Statement

I. Schiavetti has acted as a paid consultant to Associazione Commissione Difesa Vista, Eye Pharma, Hippocrates Research, and D.M.G Italia. L. Barcellini: no disclosures. C. Lapucci has received Travel grant from Novartis, Roche and Merck. F. Tazza: no disclosures. M. Cellerino reports no disclosures. E. Capello reports no disclosures. D. Franciotta received personal honoraria from Merck, Biogen, Sanofi-Genzyme, Roche. M. Inglese received grants NIH, NMSS, FISM; received fees for consultation from Roche, Genzyme, Merck, Biogen and Novartis. M. P. Sormani, received consulting fees from Merck, Biogen, Novartis, Sanofi, Roche, Geneuro, GSK, Medday, and Immunic. A. Uccelli, received grants (to his Institution) from FISM, Biogen, Roche, Alexion, Merck Serono; participated on a Data Safety Monitoring Board or Advisory Board (to his Institution) for BD, Biogen, Iqvia, Sanofi, Roche, Alexion, Bristol Myers Squibb.A. Laroni received fees for consultation from Roche, Genzyme, Merck, Biogen, Novartis, Bristol-Myers Squibb.

## Notes

### Author Declarations

The study was conducted in accordance with all national regulations and with principles of the Declaration of Helsinki and the protocol was approved by the regional ethic committee (CER Liguria: 5/2021 - DB id 11169- 21/01/2021) and at national level by AIFA/Spallanzani (Parere n 351, 2020/21).

